# One-week test-retest reliability of nine binocular tests and saccades used in concussion

**DOI:** 10.1101/19011619

**Authors:** Stephanie Long, Tibor Schuster, Russell Steele, Suzanne Leclerc, Ian Shrier

## Abstract

**Background:** Tests of binocular vision (BVTs) and ocular motility are used in concussion assessment and management.

**Purpose:** To determine the one-week test-retest reliability of 9 binocular vision tests (BVTs) and a test of saccades proposed for use in concussion management.

**Study Design:** Prospective test-retest.

**Methods:** We examined the one-week test-retest reliability of 9 BVTs in healthy participants: 3D vision (gross stereoscopic acuity), phoria at 30cm and 3m, ability of eyes to move/fixate in-sync (positive and negative fusional vergence at 30cm and 3m, near point of convergence and near point of convergence – break [i.e. double vision]) and 1 ocular motor test, saccades.

**Results:** We tested 10 males and 10 females without concussion and a mean age of 25.5 (4.1) years. The intraclass correlations suggest good reliability for phoria 3m (0.88) and gross stereoscopic acuity (0.86), and moderate reliability for phoria 30cm (0.69), near point of convergence (0.54), positive fusional vergence (0.54) and negative fusional vergence (0.66) at 30cm, and near point of convergence - break (0.64). There was poor reliability for saccades (0.34), and both positive and negative fusional vergence (0.49 and 0.43, respectively) at 3m. Limits of agreement (LoA) were best for saccade (±34%) and worst for phoria 30 cm (±121%) and ranged from ±58% to ±70% for 7 of the 8 other tests. The LoA for phoria at 3m were uninformative because measurements for 18 of 20 participants were identical.

**Conclusion:** We found test-retest reliability of the BVTs and saccades ranging from poor to good in healthy participants, with the majority being moderate.

**Clinical Relevance:** For these vision tests to be clinically useful, the effect of concussion must have a moderate to large effect on the scores of most of the tests.

**What is known about the subject:**

- Concussions may affect some parts of visual function
- 1-week test-retest reliability for most visual tests is under-studied

**What this study adds to existing knowledge:**

- We provide intra-class coefficients and limits of agreement for 10 different visual function tests commonly conducted by clinicians in patients with concussion.

## Introduction

Many brain-related disorders (e.g. concussion,^29^ Parkinson’s Disease,^6^ attention deficit hyperactivity disorder,^20^ stroke^44^) have a visual component as part of their findings.^24^ For example, posttraumatic vision impairments have been reported in 30% to 65% of patients with a mild traumatic brain injury,^11^ and are found in nearly 30% of patients with a sport-related concussion.^25^ Some symptoms associated with concussion are believed to be caused by deficits in the visual system and include: headaches, sensitivity to light, diplopia, and blurred vision.^12^

Tests of binocular vision (BVTs) and ocular motility over the last 70 years ^9^ include ^9,23,37^: gross stereoscopic acuity, near point of convergence - break (i.e. double vision), phoria at 30cm and 3m, positive and negative fusional vergence at 30cm and 3m, and saccadic eye movement assessment. Recent studies have suggested that concussion may result in deficits of convergence, binocular vision, and ocular motility.^4,12,45^ Convergence insufficiency is a disorder of binocular vision diagnosed by abnormal near point of convergence, one of many BVTs that assesses an individual’s visual capacity.

Before we can conclude that visual function is abnormal in concussion, we must first understand the test-retest reliability of vision tests in healthy control participants. Despite their frequent use, limited studies have examined the reliability of these vision tests. When evaluating the one-week test-retest reliability of the Randot Stereotest (test of gross stereoscopic acuity), 71-82% of adult and child participants with normal vision and with strabismus had perfect agreement between two measurements;^17,48^ common psychometric properties such as intraclass correlation coefficient (ICC) or limits of agreement (LoA) were not reported. An older study of near point of convergence reported an ICC of 0.65 across six testing sessions in six healthy adults, but failed to specify the number of examiners or the time interval between testing sessions.^10^ A study of near point of convergence – break in school-aged children reported excellent reliability (ICC≥0.94).^36^ A more recent study of three consecutive measurements of near point of convergence – break without a rest interval between tests, found ICCs ranging from 0.78-0.89 in young concussed athletes with convergence insufficiency and 0.92-0.97 in concussed athletes with normal vision.^31^ When examining phoria with the prism alternate cover test, one study reported 95% LoA of ±4.1 to ±7.3 prism diopters for distance and ±3.3 to ±8.3 prism diopters for near in young children with esotropia, but did not provide ICC.^32^ A one-week test-retest reliability study assessing positive fusional vergence with a prism bar reported ICCs ranging from 0.53-0.59.^36^ One to ten day test-retest reliability of the prism bar test had 95% LoA ± 4.0 for negative fusional vergence and ±13.9 for positive fusional vergence.^5^ Finally, the only study we could find that measured reliability for saccades used a computerized prosaccade task. The authors reported moderate two-month test-retest reliability (ICC=0.59) in adult participants with normal vision.^16^

The above studies provide some information regarding the reliability of these vision tests. However, vision tests are used to follow patients over time, and one might expect additional variability when patients are measured on different days due to fatigue, stress, and other factors. Understanding the usual variability that is independent of changes in pathology or recovery is essential for proper interpretation of trends in results over time. Therefore, the objective of this study was to determine the one-week test-retest reliability of 9 BVTs and a test of saccades in healthy adult participants. We evaluated versions of these visions tests that are commonly used by clinicians as opposed to versions used in research studies in order to best assess the utility of the tests in clinical practice. All tests in the present study were performed by the same clinician on two testing dates.

## Methods

### Study Design

We examined a convenience sample of healthy colleagues, friends, and social connections in Montreal, Canada on two separate occasions exactly seven days apart at approximately the same time of day (e.g. morning vs. evening). One individual clinician trained in orthoptics examined all participants individually. We arranged for 4-6 participants to be examined sequentially in a two to three-hour block of time. We randomized the order in which participants were examined using a random number generator within each block. To minimize the probability of the clinician recalling the first scores at the second visit: (1) the clinician verbally reported the results to a research assistant (recorded on paper) and was not informed of any scores until data collection was completed, and (2) at the second visit, the same group of 4-6 participants was examined over the same block of time but their examination order was changed compared to the first visit. This study was approved by the Jewish General Hospital Institutional Review Board.

### Participant Selection

We included healthy adults 18 to 35 years. There were no participants with a history of conditions that may affect BVTs (or treatment for such conditions) or saccades such as strabismus (contraindication to BVTs), migraines, neurological disorders, or currently taking muscle relaxants, selective serotonin-reuptake inhibitors, anxiolytics, stimulants, or any other drug class for other psychological conditions that might affect test results. Although there were three subjects with remote history of concussion, they had fully recovered and were not experiencing concussion symptoms or related limitations at the time of our study.

### Clinical procedures and measures

We collected the following demographic information at the first visit: date of birth, sex, highest level of education achieved, use of corrective lenses, occupation, and any relevant past medical history (e.g. migraines, vision problems, use of medication, history of concussion).

Prior to conducting the vision tests, each participant first completed the symptom portion of the validated sport concussion assessment tool (SCAT3) form.^1,51^ These results were used as a sensitivity analysis to evaluate if changes in their physical states at the two testing sessions might explain potential discrepancies.

### Vision Tests

We examined 9 BVTs and a test of saccades. For gross stereoscopic acuity, near point of convergence, near point of convergence – break and phoria, a lower score represents better vision function. For positive and negative fusional vergence, and saccades, a higher score represents better vision function. Brief descriptions of each are provided below and more details are available in the *Appendix*.

For gross stereoscopic acuity, we used the Randot Stereotest (Stereo Optical Co., Inc., Chicago, IL) according to manufacturer’s instructions.^43^ For all tests using a target, the tip of a ballpoint pen was used as the near target, and a 6cm^2^ square card mounted on a wall was used as the far target. For near point of convergence and near point of convergence - break, we followed procedures by Maples et al.^27^ The near point of convergence score was the distance (cm) between the bridge of the nose and the target at the closest point at which the individual could maintain balanced oculomotor synergy between both eyes, which is identified as when one eye diverges outwards.^9^ The near point of convergence – break score was the distance between the bridge of the nose and the point at which diplopia occurred.^7,23^ We measured phoria at 30cm and 3m using the prism alternate cover test with procedures described by the Pediatric Eye Disease Investigator Group.^32^ For positive and negative fusional vergence at 30cm and 3m, we used a horizontal prism bar (base-out for positive fusional vergence, base-in for negative fusional vergence).^19^ To evaluate saccades, we used the specific testing procedures of our clinician. In his test of saccades, participants assumed a tandem stance and attempted to move only their eyes when lights appeared and disappeared (using a gap protocol in which the first light disappeared before the second appeared) on the screen. The clinician evaluated their performance qualitatively along three measures: quality (bad, medium, good), synchronization (bad, medium, good), and saccadic correction (many, few, none).

## Statistical Analysis

### Reliability Estimates

We evaluated test-retest reliability using statistical measures for consistency and accuracy i.e. the intra class correlation coefficient (ICC)^42^ and 95% limits of agreement (LoA).^8^ We considered an ICC of ≤0.5 as poor, 0.51 - 0.74 as moderate, 0.75 - 0.89 as good, and ≥0.90 as excellent reliability.^26^ When multiple participant scores were the same, we used the jitter function in the R software^34^ to slightly modify the scores when plotting the results so they would appear distinct from each other.

The LoA were calculated as recommended in the units of the scale measured.^8^ To compare LoA across tests, we also standardized the scores and reported them as percentage difference [(T1-T2)/mean(T1&T2)*100]. The LoA results were summarized graphically with Bland-Altman plots.^8^ We used the raw scale measures commonly known to clinicians for the y-axis, and report the standardized version in parentheses to provide an overview of all tests.

Due to the limited sample size and to avoid being overly conservative in our evaluation, we followed the practical solution for addressing multiple testing proposed by Saville.^38^ Formal multiplicity correction of confidence levels was not performed but we thoroughly report all statistical assessments enabling an informal *type I error* assessment by the reader.

### Effect of Physical State

As a sensitivity analysis, we wanted to explore if a participant’s change in physical state could have been associated with their vision test scores. We, therefore, compared the change in BVTs and saccade scores against the change in SCAT3 symptom scores using the Pearson’s correlation coefficient. Although we report *p*-values for these comparisons, we caution that these are minimum values as we did not correct for multiple testing.

All statistical analyses were performed on R software 3.4.3.,^34^ plots were created using the ggplot2 package.^50^

### A priori sample size calculation

Sample size calculations were done prior to the study. We used a precision-based method for sample size calculations based on the ICC. Since the expected estimate for ICCs is zero, our sample size calculation relied on specifying the maximum acceptable width of the confidence interval for the measure of agreement. We considered the lower bound of clinical acceptability to be an ICC of 0.5,^49^ and expected the true ICC for repeated assessment of the different vision test scores to be at least 0.75. Therefore, we believed that our estimate imprecision (95% confidence interval width / 2) should not exceed 0.25. With 20 participants, under the postulated assumptions, the precision of the ICC estimate was anticipated to be ± 0.20.^49^

## Results

Of 47 potential participants identified, 26 had scheduling conflicts, one was not able to attend the second visit and was excluded from the analysis, leaving 20 participants for the final analysis.

Demographic data for the sample analyzed is included in Table 1. Our sample included 50% females with an average age of 25.5 years (SD=4.1, range=18-35), almost all university educated, with 55% wearing corrective lenses with an up-to-date prescription. None of the participants had any history of vision symptoms or a history of past binocular vision therapy. There were six participants who had previously sustained one or tws concussions, which occurred 2 to 15 years prior to our study; none of these individuals reported any residual concussion symptoms at the time of our study. Additionally, the three participants who had reported taking selective serotonin reuptake inhibitors or anxiolytics had not received the medication for at least five years as they no longer suffered from the condition.

**Table 1.**
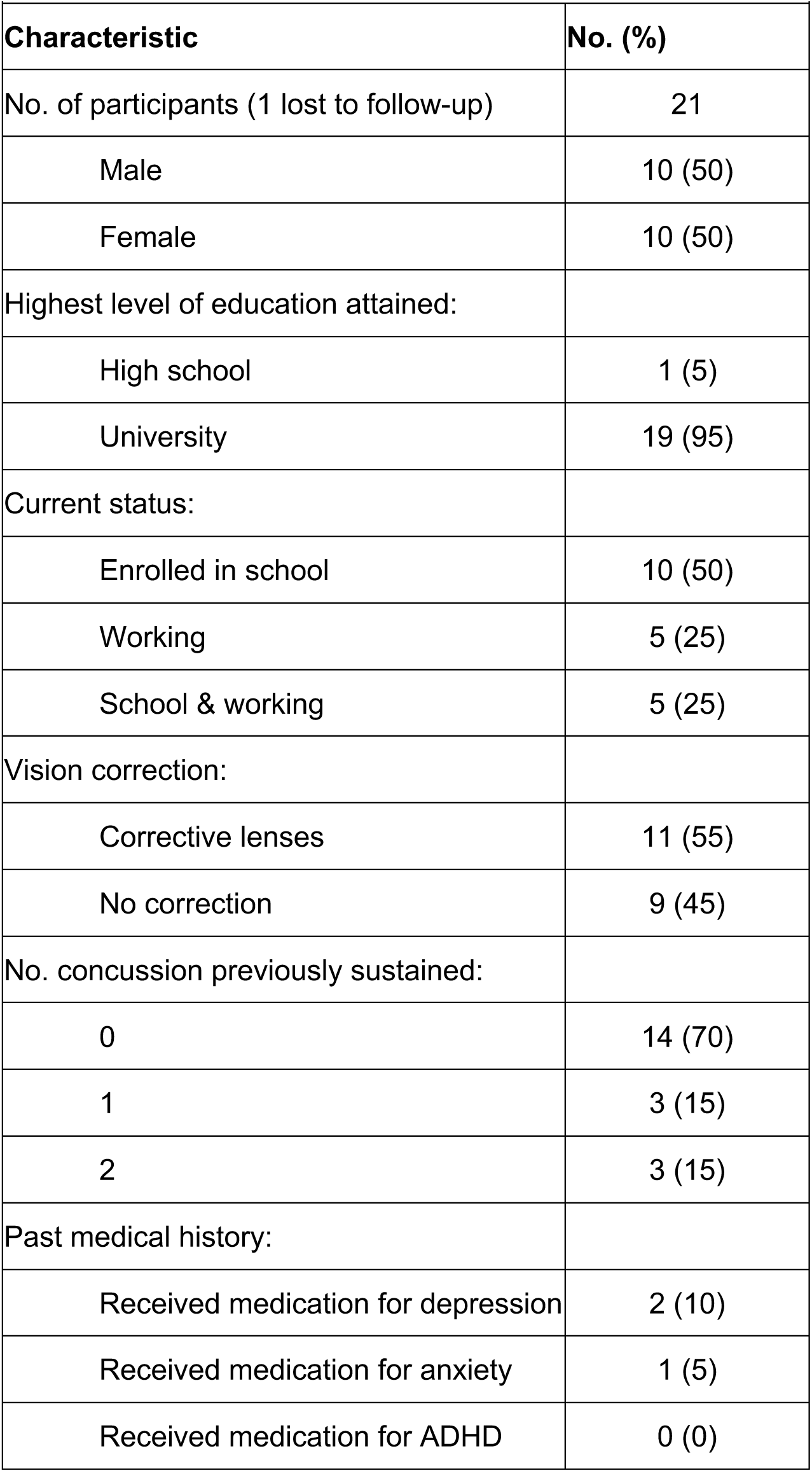
Demographic and clinical characteristics of participants.

| **Table 1: Demographic and clinical characteristics of participants** |  |
| --- | --- |
| **Characteristic** | **No. (%)** |
| No. of participants (1 lost to follow-up) | 21 |
| Male | 10 (50) |
| Female | 10 (50) |
| Highest level of education attained: |  |
| High school | 1 (5) |
| University | 19 (95) |
| Current status: |  |
| Enrolled in school | 10 (50) |
| Working | 5 (25) |
| School & working | 5 (25) |
| Vision correction: |  |
| Corrective lenses | 11 (55) |
| No correction | 9 (45) |
| No. concussion previously sustained: |  |
| 0 | 14 (70) |
| 1 | 3 (15) |
| 2 | 3 (15) |
| Past medical history: |  |
| Received medication for depression | 2 (10) |
| Received medication for anxiety | 1 (5) |
| Received medication for ADHD | 0 (0) |

Baseline (i.e. Visit 1) scores for all tests quantitatively scored can be found in Table 2. The mean scores of these tests are within range of normative scores reported in the literature (see *Appendix* for more details).

**Table 2.** Baseline scores for quantitatively scored vision tests.

| **Table 2: Baseline scores for quantitatively scored vision tests** |  |
| --- | --- |
| **Test (normal range)** | **Mean (SD)** |
| Gross stereoscopic acuity (20-100 arc seconds) | 42.5 (20.2) |
| Near point of convergence (3-7 cm) | 4.6 (1.1) |
| Near point of convergence – break (1-8 cm) | 5.2 (1.5) |
| Phoria – 3m (44-90 prism diopters) | 0.3 (1.0) |
| Phoria – 30cm (1-45 prism diopters) | 19.5 (11.2) |
| Positive fusional vergence – 3m (6-45 prism diopters) | 16.8 (8.5) |
| Positive fusional vergence – 30cm (10-40 prism diopters) | 25.3 (9.5) |
| Negative fusional vergence – 3m (2-8 prism diopters) | 5.4 (1.8) |
| Negative fusional vergence – 30cm (4-30 prism diopters) | 18.5 (6.7) |

Two BVTs achieved good reliability. For phoria 3m (ICC=0.88), 18 out of 20 pairs of measurements were identical at test and retest, with one participant scoring 0 and 1 prism diopters, and the other scoring 0 and 2 prism diopters. Due to this highly skewed score distribution, the 95% LoA are relatively uninformative (data not shown). The ICC for gross stereoscopic acuity was 0.86 (Figure 1). For this test, 5 out of 20 pairs of measurements were identical, with the remaining pairs differing by 5 to 40 arc seconds; the 95% LoA was ±27.6 arc seconds.

**Figure 1:**
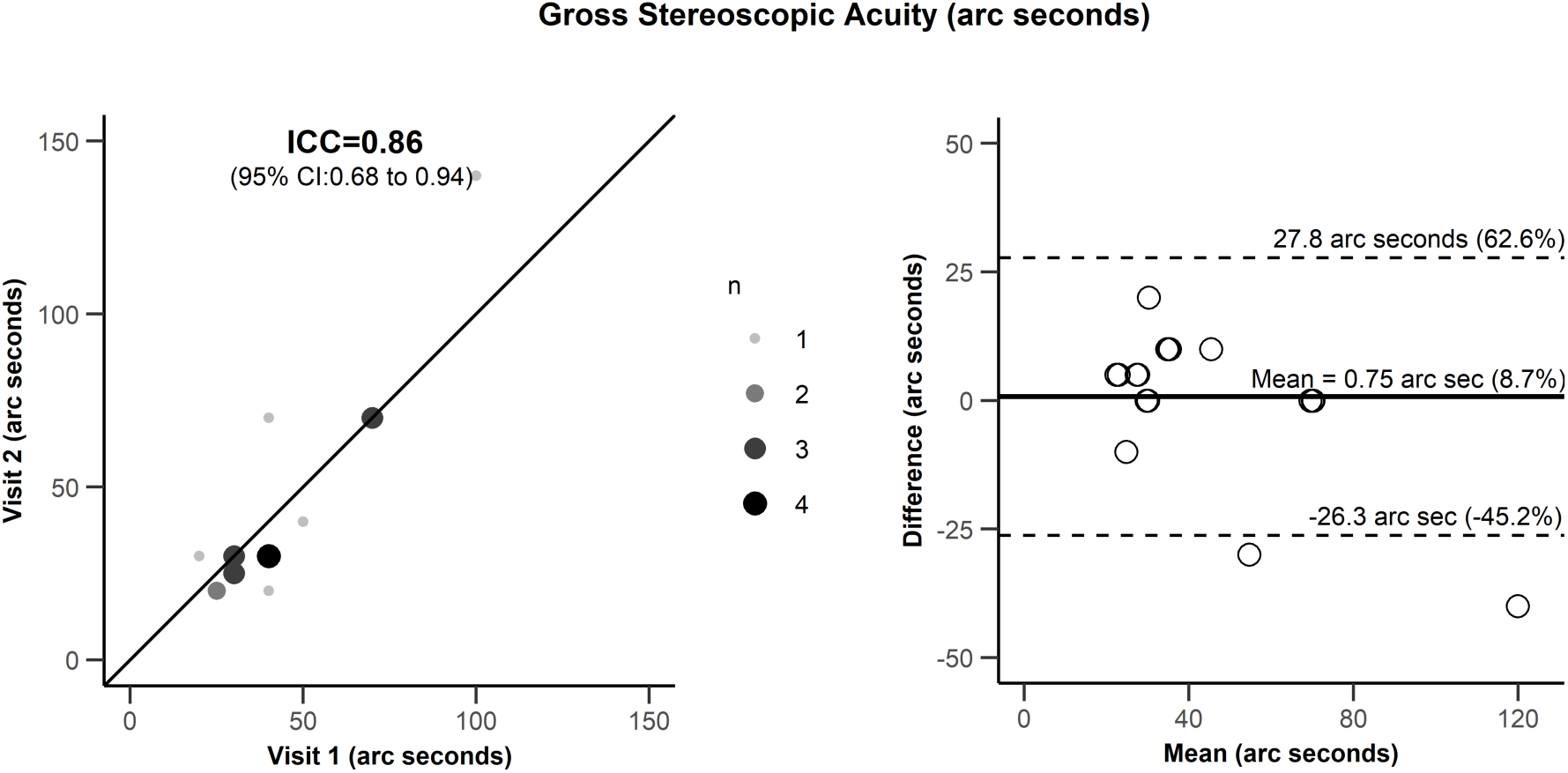
The left graph is a scatter plot for the test results at the first visit (x-axis) and retest results at the second visit (y-values) for gross stereoscopic acuity (GSA). The intra-class coefficient and its 95% confidence intervals are illustrated on the graph. The size of the gray dots (and n in the legend) represents the number of subjects with the values shown on the graph. The line of equality indicates where all points would fall if reliability was perfect. The right graph represents the Bland-Altman plot with the mean of the test-retest values on the x-axis and the difference between the test-retest results on the y-axis. The solid line represents the bias and the dotted lines represent the 95% limits of agreement (LoA). The y-axis scale represents the raw units of the test because these are the most relevant to the clinician treating the patient. Because we conducted many tests and readers may be interested in comparing the LoA across tests, we also report LoA as percent difference (T1-T2/mean of T1&T2) in parentheses.

We found moderate reliability for near point of convergence, near point of convergence – break, phoria 30cm, and for positive and negative fusional vergence at 30cm (Figures 2 and 3), with ICCs ranging between 0.54 to 0.69. For these BVTs, the 95% LoA was ±2.5 cm for near point of convergence, ±2.5 cm for near point of convergence – break, ±16.3 prism diopters for phoria 30cm, and ±17.3 prism diopters and ±10.4 prism diopters respectively for positive and negative fusional vergence at 30cm.

**Figure 2:**
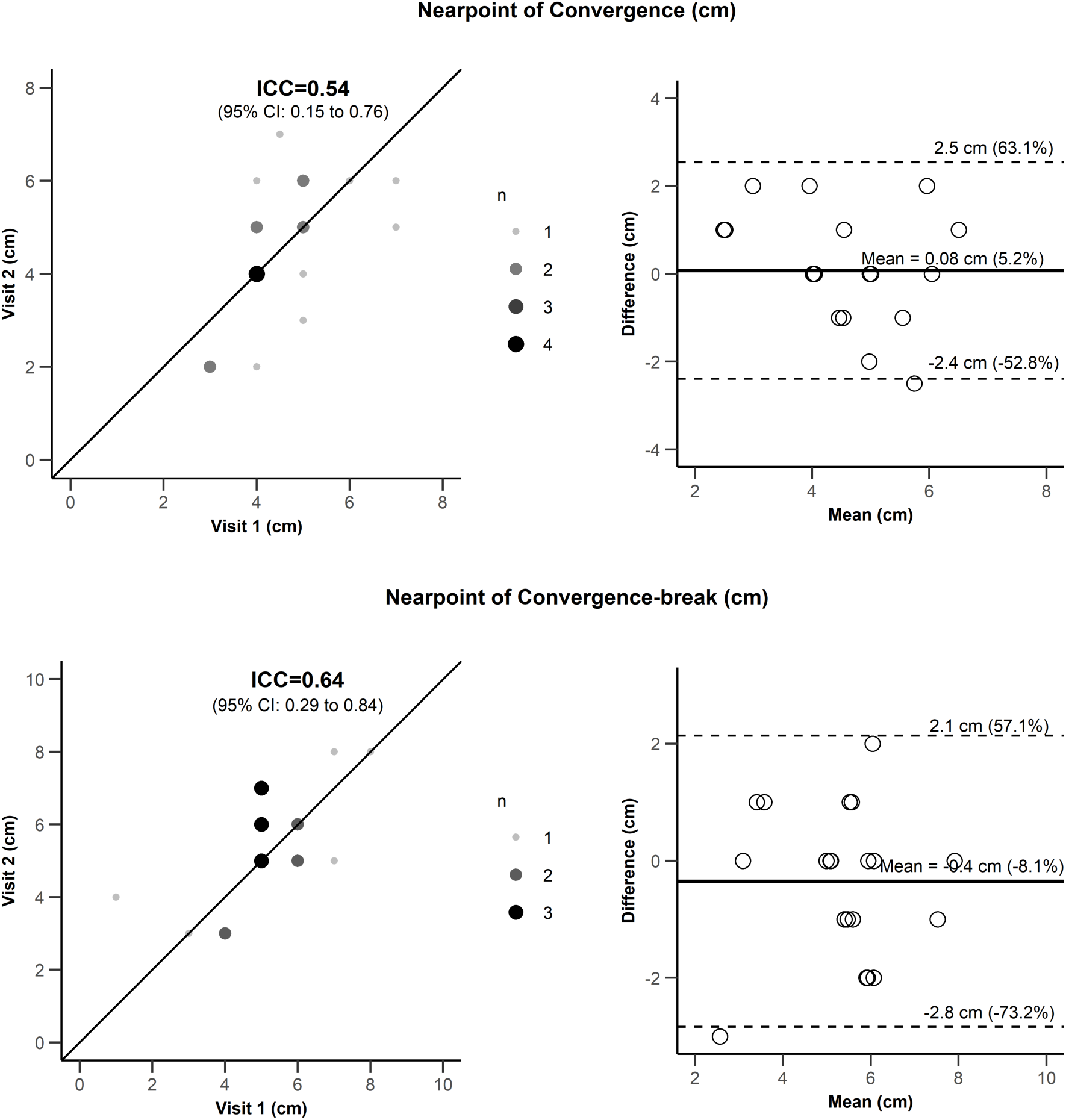
Scatter plots with intra-class coefficient results for the test-retest results (left) and limits of agreement (LoA, right) for one-week test-retest reliability for 2/5 binocular vision tests with moderate reliability. Legends are identical to Figure 1.

**Figure 3:**
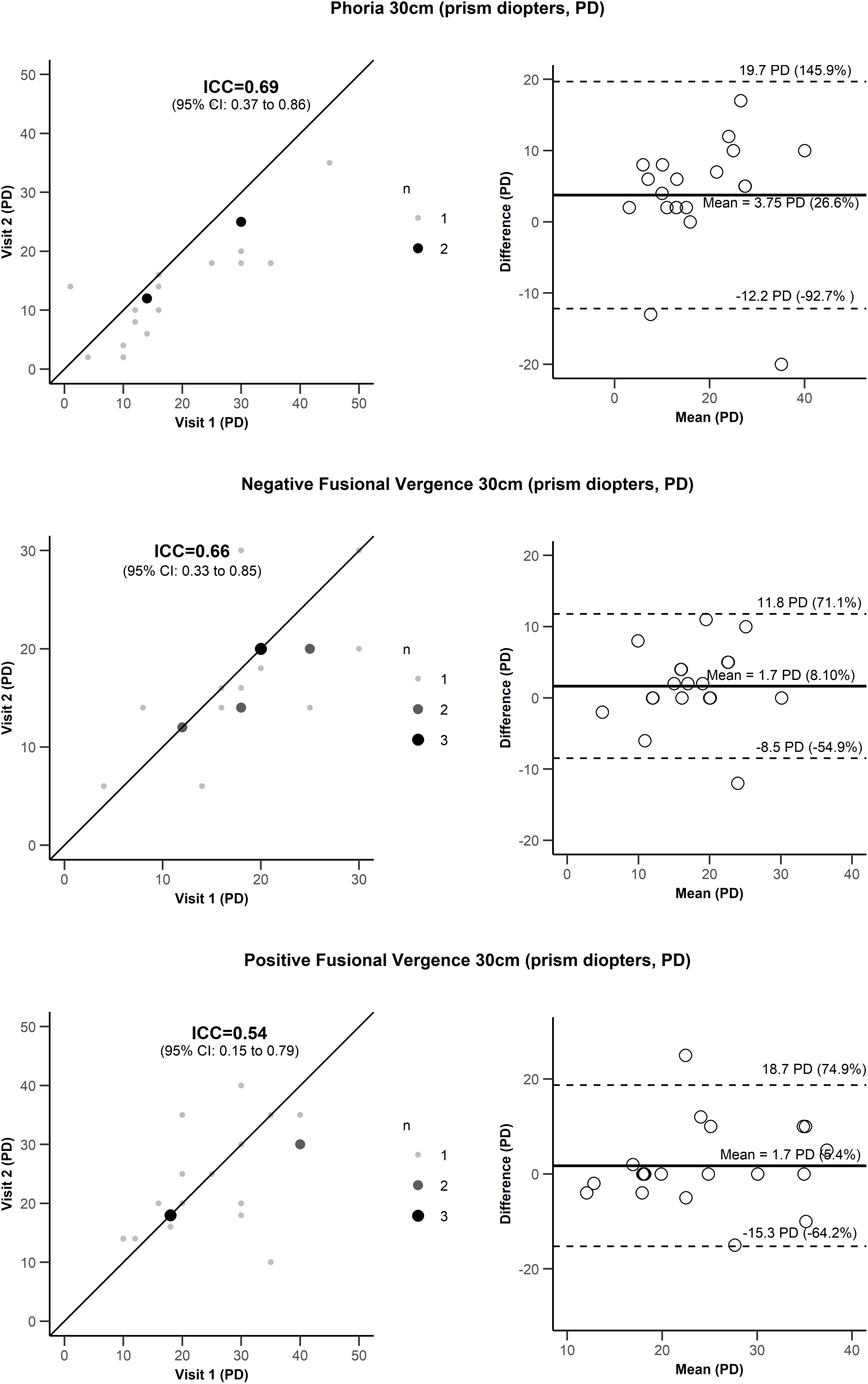
Scatter plots with intra-class coefficient results for the test-retest results (left) and limits of agreement (LoA, right) for one-week test-retest reliability for the remaining 3/5 binocular vision tests with moderate reliability. Legends are identical to Figure 1.

The three tests with poor reliability were positive and negative fusional vergence at 3m, and saccades (Figure 4).

**Figure 4:**
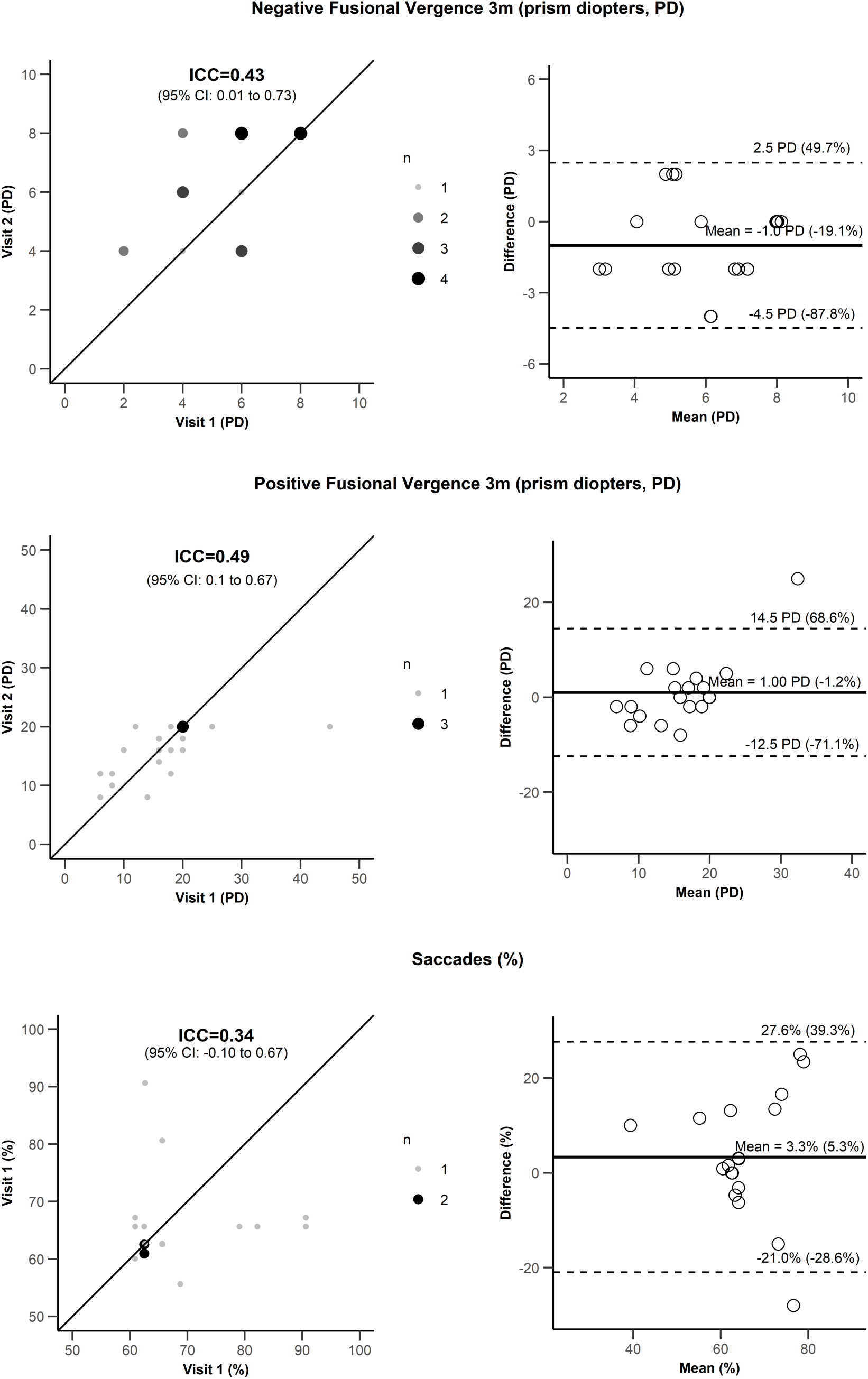
Scatter plots with intra-class coefficient results for the test-retest results (left) and limits of agreement (LoA, right) for one-week test-retest reliability for saccades and the 2 binocular vision tests with poor reliability. Legends are identical to Figure 1.

### Effect of Changes in Physical State

In our sensitivity analysis, a participant’s physical state (as measured by the SCAT3 symptom score) was not relevantly associated with their BVT scores across testing sessions. Pearson’s correlations between changes in vision test scores and changes in symptom scores ranged from −0.006 to 0.31 for all vision tests (*p*-values ranged from 0.19 to 0.98).

## Discussion

Our results suggest that only 2 out of 10 vision tests demonstrated good reliability, and 5 additional tests had moderate reliability. There was poor reliability for saccades and both positive and negative fusional vergence at 3m. The 95% LoA suggests that even with good or moderate reliability, one can expect that scores for an individual with repeated measures may vary by 50-70% of the mean score across all measures even if there is no change in visual function. These results highlight the need for more accurate, quantifiable, and repeatable tests since one might expect even more variability in a patient population compared to the healthy population that we studied. Further studies are necessary to determine if changes to visual function with concussion or other neurological injury (the resultant signal) are large enough to be noticed given the amount of inherent noise in the tests.

We used the Randot Stereotest to assess gross stereoscopic acuity. Our ICC of 0.86 and 95% LoA of ±27.6 arc seconds support previous findings of good reliability in adults in the one-week time frame. The 95% LoA were previously reported as ±0.57 *log arc seconds* (our results are ±0.58 log arc seconds using the same method of calculation) based on 36 patients between 7 to 76 years of age; time between testing intervals ranged from 10 to 364 days using a different examiner at each time point.^2^ Another study reported that 82.0% of their participants had identical results at test and retest taken same day in 111 adult and children with normal vision, but normal psychometric properties such as ICC or LoA were not reported.^48^ A study examining the one-week test-retest reliability of gross stereoscopic acuity using the related Titmus fly test in 90 children reported perfect reliability (ICC=1.0)^28^. The Random dot “E” stereotest reported only interrater agreement (*Kw*=0.33-0.44) in 1257 children, but not test-retest reliability.^46^

In the literature, there is no clear distinction between measures of near point of convergence and near point of convergence – break. Further, near point of convergence is sometimes referred to as a measurement of gross convergence with fusional convergence^22^ and other times as gross convergence with proximal convergence.^21,22,47^ The inconsistent use of these terms complicates comparisons across studies when the measurement procedure is not reported. The Convergence Insufficiency Treatment Trial Study^13^ considered near point of convergence – break as the point “[w]hen diploplia was reported,” which is consistent with our definition.^7,23^ We defined near point of convergence as the closest point at which one eye diverges outwards.^9,23,37^ Across the literature, near point of convergence and near point of convergence – break are sometimes used interchangeably. For instance, near point of convergence has been defined as the point “when the target blurs, jumps or becomes double,”^37^ “when [the participant] saw 2 distinct images,”^31^ and “when the patient reported diplopia”.^4^

We found moderate reliability for both near point of convergence and near point of convergence – break, whereas others have reported good to excellent reliability.^31,36^ In our measurement procedure, participants fixated on a target that the clinician moved towards their eyes in free space as used by some clinicians and researchers in the concussion field.^29^ Others used an accommodative target, such as the Royal Air Force (RAF) rule,^7,37^ or Astron International (ACR/21) Accommodative Rule.^13,22,36,40^ The RAF rule had good (ICC=0.84) test-retest reliability for near point of convergence in 3 subjects with idiopathic neck pain and 7 healthy subjects, but the test interval was only specified as less than one-week.^18^ The Astron International Accommodative Rule had excellent one-week test-retest reliability (ICC=0.94-0.98) in 20 healthy children for near point of convergence – break.^36^ Although tests using these accommodative targets may have increased reliability compared to the methods used in this study, one would generally like to minimize the accommodative load in patients with concussion because it may increase symptoms. Some of the variability in our results is likely explained by accommodation variability in our participants. We found no studies directly comparing the reliability of the different procedures. Sheiman et al ^39^ and Rouse et al ^36^ provide a more complete discussion of the advantages and disadvantages of the different methods used to assess near point of convergence.

Measurements of phoria using the prism alternate cover test had good reliability for distance (ICC=0.88) and moderate reliability for near (ICC=0.69). These results are consistent with other studies which measured adult and child participants with strabismus or esotropia,^14,32^ even though none of our participants had these conditions. Despite the similarity in findings, our analysis methods differed slightly. For instance, because different prism increments are used to measure smaller (2-20 prism diopters) or larger (>20 prism diopters) angles, other authors analyzed and reported these strata separately.^14,32^ Unlike other authors, we evaluated all angles of deviation together.

For both positive and negative fusional vergence, we reported moderate reliability (ICC=0.54, 0.66 respectively) for near fixation and poor reliability (ICC=0.49, 0.43 respectively) for distance fixation. These results are contrary to studies reporting lower within-subject variability at near fixation,^33^ or no differences due to distance.^5^ Further, we found that negative fusional vergence had slightly higher reliability than positive fusional vergence at near, but were less reliable at distance. However, standard clinical practice and evidence suggests the opposite; negative fusional vergence is considered to have less reliability than positive fusional vergence.^3,35^ It is possible that the order in which fusional vergences are taken may influence their scores. We measured fusional vergences grouped by distance: (1) negative fusional vergence far, (2) positive fusional vergence far, (3) negative fusional vergence near, and (4) positive fusional vergence near. However, the reliability was poor for both tests at distance suggesting the order of test administration would not explain the discrepancy between our results and the literature. It remains possible that our results are different than others because of slight differences in our methods that are not apparent in the description of the tests (see *Appendix* for full description of our methods).

The test of saccades had the lowest ICC and poorest reliability of all the vision tests. We used the clinical procedures our clinician uses in his daily practice with his patients. Participants assumed a tandem stance and attempted to follow appearing and disappearing lights on a screen under a gap paradigm with only their eyes, trying to keep their head still. The clinician stood beside the screen in front of the participant to observe their eye movements. In some published saccade test protocols, the participant’s head is held still with a chin rest and a forehead support to ensure that only the eyes are tracking the movements.^16,30^ In the NSUCO oculomotor test, the head is not held still.^41^ However, unlike other tests of saccades, our clinician had patients take a tandem stance which adds an additional vestibular challenge. The added challenge may influence their performance on the task and introduce more noise. Finally, the evaluation of the saccades was qualitative, dependent solely on the judgment of the clinician with no objective measure.

### Strengths and Limitations

We selected a seven-day interval between testing times to evaluate the test-retest reliability of the vision tests. This allowed for normal variation over time due to sleep, stress, and other factors in order to provide an ICC that is applicable to following patients over time. In addition, it avoids any lingering symptoms following a test that might lead to an underestimated ICC. The seven-day interval also increased the likelihood the clinician remained blinded to the previous results and facilitated participant recruitment because we could select a day and time when participants were generally available. Some studies previously evaluated interrater reliability.^14,17,32,36^ Although this has merit when one is interested in tests being evaluated by more than one clinician as what might occur in group practice or a research study, interrater reliability is less important when patients are followed by a single clinician over time. Our objective was to define the expected “noise” when a single clinician follows a single patient over time, as would occur in our target condition (i.e. concussions), so that clinicians can appropriately interpret changes in the vision test scores. We provided results based on different perspectives of reliability. The ICC is a measure of variability due to genuine differences in the participant or due to measurement error. For instance, the ICC was 0.88 for phoria 3m, indicating that 88% of the variability in the measurements was due to differences between participants, and 12% was due to noise within the measurement of a participant. In addition, the 95% LoA provides the magnitude of the noise that can be expected with repeated measures. Differences between tests at baseline and after diagnosis of a condition (e.g. concussion) likely represent a true signal of a change in vision tests within the patient if these differences are larger than the noise (i.e. LoA) found in our study.

Our study also had limitations with respect to participant population and testing measures. We had a relatively small homogeneous sample size of 20 participants who were recruited via convenience sampling in a university setting. However, our participants did include an equal number of males and females, over half wore corrective lenses, and the age range was 18 to 35 years. Our study population was thus relatively representative of our target population of young athletes who may sustain concussions. The approaches used by our clinician were standard to his clinical practice and were used on both of the testing days for all participants. However, the methods he used to assess vision function sometimes differed from testing procedures reported in the literature. For instance, a testing distance of 30 cm was used instead of the standard 40 cm distance for near testing of positive and negative fusional vergence.^5^ The near testing distance of 30 cm was also used for phoria, which is similar to the distance commonly used in the literature, 1/3 m.^14,32^ However, we are not aware of any studies comparing the effect of distance on reliability. Our clinician did not attempt to separate out accommodative testing from convergence (i.e. near point of convergence and near point of convergence - break, also known as relative convergence) although this may be possible.^15^ Therefore, our measure of convergence could have been affected by accommodative issues. The saccadic eye movement test of our clinician also differs from commonly used tests in clinical practice and the scoring of this test was qualitative and subjective, which could lead to increased variability and inconsistency in scoring. Developing more quantifiable and reliable testing methods is particularly important for conditions such as concussions, as they are characterized by many symptoms which may only lead to subtle changes that are not detectable with imprecise tests.

## Conclusion

We found that only 2 of 9 BVTs had good one-week test-retest reliability that could detect small to moderate changes in visual function, and an additional 5 BVTs that might be able to detect moderate change in visual function. The remaining two BVTs and saccades may still be useful if changes in visual function are expected to be larger than the noise of the measure.

## Data Availability

The raw data on the test-retest scores used in these analyses is available on the Open Science Foundation (https://osf.io/nbskx/wiki/home/).

https://osf.io/qzfj8/

## Acknowledgements

We would like to thank Isabel Pereira for her help throughout the course of this work. We would also like to thank David Tinjust, the clinician who conducted these tests in our participant population and who provided partial funding for this study. This study was funded through programs designed to foster collaboration between academics and industry. The government sources are the MITACS program, and the MEDTEQ program. The industry partners were Apexk Inc, Varitron Technologies Inc and l’Institut National du Sport du Quebec.

## Appendix

The Appendix expands on the concise description of the nine binocular vision tests and test of saccades provided in the manuscript. This is especially important when comparing results across different studies in evidence synthesis.

## Notes

### Competing Interest Statement

The authors have declared no competing interest.

